# Tuberculosis diagnosis cascade in Blantyre, Malawi: a prospective cohort study

**DOI:** 10.1101/2020.06.16.20132472

**Authors:** Helena R A Feasey, Elizabeth L Corbett, Marriott Nliwasa, Luke Mair, Titus H Divala, Wala Kamchedzera, McEwen Khundi, Helen E D Burchett, Emily L Webb, Hendramoorthy Maheswaran, S Bertel Squire, Peter MacPherson

**Author notes:** **Corresponding author:** Helena Feasey, London School of Hygiene and Tropical Medicine, Keppel Street, Bloomsbury, London, WC1E 7HT, UK.

## Abstract

**Background:** Tuberculosis (TB) control relies on early diagnosis and treatment. International guidelines recommend systematic TB screening at health facilities in high-burden settings, but implementation is challenging. To construct a TB diagnosis care cascade, we investigated screening guideline adherence and completion of TB testing steps in Blantyre, Malawi.

**Methods:** A prospective cohort recruited adult (≥18) outpatients attending Bangwe acute-care primary clinic between 21/5/2018 and 6/9/2018. Entry interviews recording TB symptoms and demographic characteristics were linked to exit interviews by biometrics. Care cascades were constructed to estimate the proportion of patients progressing through each step of the diagnostic pathway. Multivariable logistic regression was used to investigate factors associated with being asked to submit sputum.

**Results:** Of 5,442 clinic attendances 2,397 (44%) had exit interviews. In clinically indicated participants (n=330) 203 (61.5%) were asked about cough, 39 (11.8%) were asked for sputum, 27 (8.2%) gave sputum and 1 (0.3%) received same-day results. Significant associations with request for sputum were: any TB symptom (aOR:3.20, 95%CI:2.02-5.06), increasing age (aOR:1.02, 95%CI:1.01-1.04 per year) and for HIV-negative participants only, a history of previous TB (aOR:3.37, 95%CI:1.45-7.81). Numbers requiring sputum survey (20/day) outnumbered diagnostic capacity (8-12/day).

**Conclusion:** Patients were lost at every stage of the TB care cascade, with same day sputum submission achieved in only 8.2% if clinically indicated. Infection control strategies should be implemented, with reporting on early steps of the TB care cascade formalised. High-throughput interventions, such as digital CXR, that can optimise same-day TB screening are urgently needed to meet WHO End TB goals.

**Summary:** *What is already known?:*

- WHO guidelines recommend systematic screening for Tuberculosis at health facilities in high-burden settings, but implementation is challenging.
- Care cascades have been widely used by HIV programmes to evaluate care delivery but have only recently been applied to TB care. Care cascades help to define the steps of the cascade most in need of intervention

*What are the new findings?:*

- In this study, only 8.2% of those clinically indicated to test for TB (as per national guidelines in Malawi) did so, with patients lost at every step of the diagnosis care cascade.
- Failure to request sputum by clinicians despite elicited symptoms led to the biggest single gap in the diagnosis care cascade, followed by not asking about symptoms.
- If all patients attending the clinic were screened for TB as per the guidelines, the current testing facilities would only be able to process up to two thirds of the required samples.

*What do the new findings imply?:*

- Interventions focusing on health worker behaviour may have the greatest potential for retaining presumptive TB patients within the diagnosis cascade
- We must formalise and strengthen reporting on the early steps in the TB care cascade: a requirement to report numbers of screened presumptive TB cases would allow greater focus on these critical steps.
- If identification of presumptive TB patients is subsequently improved a novel high-throughput approach to triage testing using new diagnostics will be required for LMICs to increase capacity.

## Introduction

Tuberculosis (TB) is the leading infectious cause of death worldwide and an estimated 10 million people developed TB disease in 2018 (1). TB control relies on early diagnosis and treatment, as reflected in the World Health Organization (WHO) End TB 2025 target of ≥90% of people who develop TB being notified and treated (2). To achieve this the WHO recommends systematic TB screening for priority risk groups in order to reduce poor disease outcomes and TB transmission (3). These recommendations are reflected in many National TB Programme (NTP) guidelines (4-7).

The TB care cascade is “a model for evaluating patient retention across sequential stages of care required to achieve a successful treatment outcome” (8) that quantifies gaps in care delivery and adherence to guidelines. Care cascades have been extensively used to evaluate HIV care delivery (9), but have only recently been applied by TB programmes (8) to expand analysis beyond standardised treatment outcome reporting (10) and adhoc diagnostic pathway analysis (11).

Subbaraman et al’s generic model for a care cascade for active TB identifies the first gap as “did not access a TB diagnostic test” (8). This first gap, is repeatedly the largest in many settings (12, 13), in keeping with the numerous issues relating to sputum-based tests (14).

Recent studies have emphasised variability in TB diagnosis cascades in high burden countries. In India, only 12-17% of patients were correctly asked to test for TB (15), whereas in Nairobi, Kenya (16) completion was 50% and a recent systematic review found a range from 4% in Thailand to 84% in South Africa (17). These follow earlier studies (2011-12) where provider request for sputum was 35% in Botswana (18) and 21% in Uganda (19). Unlike HIV programmes, which have widely adopted the cascade approach (9, 20), most high-burden TB countries, do not routinely collect data to estimate adherence to systematic TB screening guidelines in health facilities.

WHO recommends that “people living with HIV should be systematically screened for active TB at each visit to a health facility” and that “systematic screening for active TB should be considered among people who are seeking health care… and who belong to selected risk groups” high-burden TB settings (3). These risk groups include older people and those previously treated for TB. However, an estimated 36% of new TB cases are still not identified or officially notified, partly due to failure to diagnose active TB in people accessing healthcare (21). Examining TB test access for these risk groups and subsequent steps in the TB diagnosis cascade will be critical for efficient TB programme design.

The aims of this study were to: construct a TB diagnosis care cascade; describe the proportion of “clinically-indicated” patients (defined by the Malawi National guidelines (4)) who progressed through each step of the diagnosis cascade in a primary care clinic; and investigate factors associated with being offered a TB test.

## Methods

### Study Design

A prospective cohort of adults aged 18 and over was recruited from May to September 2018. The study formed part of the pilot phase of a pragmatic randomised trial at Bangwe health clinic in Blantyre, Malawi (22).

### Study site and population

Patients self-presenting to free-of-charge acute-care services in Bangwe Health Centre – a government primary care clinic – were recruited prospectively. There are no physicians at the clinic; care is provided by nurses and clinical officers. There is a GeneXpert machine for TB sputum diagnosis and TB treatment is available on site.

Malawi National TB Programme guidelines stated that all HIV-positive adults presenting to healthcare facilities with any TB symptom (any of cough, night sweats, fever or weight loss) should receive a sputum test for TB (4). For HIV-negative adults sputum tests are recommended for all those with TB symptoms of two weeks or more.

### Data collection

Research assistants stationed at the registration desk in the acute-care clinic asked all patients for verbal consent to participate. A fingerprint scan with demographic details was recorded electronically at entry interview. Additional research assistants positioned by the two clinic exits asked all adults leaving the clinic to participate in exit interviews. Participants provided written or witnessed fingerprint (if illiterate) consent for exit interviews.

Entry and exit interviews were linked through digital fingerprint bio-identification. Entry interviews recorded age, sex and presence and duration of TB symptoms. Exit interviews asked about care received at the clinic and included self-reported HIV status and previous TB diagnosis; whether a health worker had enquired about cough; if they had been asked to submit sputum; if they submitted sputum; and if sputum results had been received. Questionnaires were kept brief to minimise inconvenience and maximise the completeness of capture.

### Statistical methods

Summary statistics compared characteristics (collected at clinic entry) of participants who had exit interviews with those who had not. Participant characteristics were also compared by HIV status (HIV-positive, HIV-negative, status unknown/never tested). “Chronic cough” was defined as cough ≥2 weeks. “Any TB symptom” included any reported cough, fever, weight loss or night sweats (23).

Diagnosis care cascades were constructed based on all participants, and separately for clinically-indicated groups: HIV-negative participants with chronic cough and people living with HIV (PLHIV) with any TB symptom. Generic care cascade Step 2 ‘Accessed TB tests’ (8) was expanded to explore symptom enquiry (cough); request to submit sputum; and sputum submission.

Univariable and multivariable logistic regression were used to investigate associations of clinical and demographic characteristics with request for sputum submission. Those who reported being on TB treatment (77 people) or isoniazid preventive therapy (IPT) (71 people) at clinic entry were removed from final analysis.

### Ethical considerations

Approval was received from the research ethics committees of the College of Medicine, Malawi and Liverpool School of Tropical Medicine. All participants provided written informed consent (or witnessed, thumb-print consent if illiterate).

### Data and reproducibility

Data and code to reproduce this analysis is available from https://github.com/petermacp/tbcascade.

### Patient and public involvement (PPI)

Public involvement included community meetings describing the study, conducted with traditional leaders and community advisory groups in Bangwe, and with health workers and patient groups at Bangwe health centre. The community advisory group will help form the dissemination plan for reporting study findings.

## Results

### Clinic attendee characteristics

Of 5,442 clinic attendances 2,397 (44%) had matched exit interviews, mainly reflecting limited study capacity to interview everyone leaving the clinic. Five individuals declined to participate in entry interviews and were not included in the study. None refused to participate in exit interviews.

Participants with matched exit interviews had similar characteristics to those with just an entry interview, with the exceptions that men were more likely to complete an exit interview (46.5% vs 42.9%, p=0.01) as were those with any TB symptom (45.4% vs 42.7%, p=0.04).

### Exit interviewee characteristics

Of the 2,397 with matched exit interviews 900 (37.5%) were male. Median age was 28 years (range 18 – 89) (Table 1). A total of 849 (35.4%) had a cough, with 221 (9.2%) having chronic cough, and 1,370 (57.2%) having any TB symptom. Previous TB treatment was reported by 141 (5.9%). Among HIV positive participants (292, 12.2%) almost all were taking anti-retroviral therapy (ART) (276, 94.5%). Of those completing exit interviews 1,485 (62.0%) self-reported good health.

**Table 1:** Baseline characteristics of exit interview participants by HIV status.

|  | HIV+<br>(N=292) | HIV- (N=1809) | Don't<br>know/never<br>tested<br>(N=296) | P value |
| --- | --- | --- | --- | --- |
| Sex (Female) | 213 (72.9%) | 1,134 (62.7%) | 150 (50.7%) | <0.001 |
| Age Median<br>(Range) | 36 (18-70) | 27 (18-87) | 27.5 (18-89) | <0.001 |
| Age 18-29 | 89 (30.5%) | 1,031 (57.0%) | 159 (53.7%) | <0.001 |
| 30-39 | 98 (33.6%) | 391 (21.6%) | 46 (15.5%) |  |
| 40-49 | 68 (23.3%) | 190 (10.5%) | 25 (8.4%) |  |
| 50-59 | 22 (7.5%) | 95 (5.3%) | 27 (9.1%) |  |
| 60-89 | 15 (5.1%) | 102 (5.6%) | 39 (13.2%) |  |
| Cough | 121 (41.4%) | 618 (34.2%) | 110 (37.2%) | 0.044 |
| Cough days (if<br>cough) Median<br>(Range) | 7 (1-3650) | 4 (1-2190) | 4 (2-1095) | 0.001 |
| - On TB<br>treatment* | 18 (14.9%) | 20 (3.2%) | 2 (1.8%) | <0.001 |
| - TB treatment<br>last 6 month* | 2 (1.7%) | 7 (1.1%) | 0 (0.0%) | 0.446 |
| - On IPT* | 26 (21.5%) | 7 (1.1%) | 2 (1.8%) | <0.001 |
| Weight loss | 65 (22.3%) | 223 (12.3%) | 28 (9.5%) | <0.001 |
| Fever | 92 (31.5%) | 550 (30.4%) | 92 (31.1%) | 0.915 |
| Night sweats | 56 (19.2%) | 342 (18.9%) | 63 (21.3%) | 0.629 |
| Any symptoms<sup>†</sup> | 181 (62.0%) | 1,007 (55.7%) | 182 (61.5%) | 0.035 |
| Chronic cough<sup>‡</sup> | 44 (15.1%) | 149 (8.2%) | 28 (9.5%) | 0.001 |
| Previous TB | 66 (22.6%) | 66 (3.6%) | 9 (3.0%) | <0.001 |
| ART | 276 (94.5%) | 0 (0%) | 0 (0%) | <0.001 |
| Self-reported<br>general health |  |  |  |  |
| Very Good | 5 (1.7%) | 58 (3.2%) | 13 (4.4%) | 0.062 |
| Good | 159 (54.5%) | 1,091 (60.3%) | 159 (53.7%) |  |
| Fair | 122 (41.8%) | 622 (34.4%) | 119 (40.2%) |  |
| Poor/Very<br>poor | 6 (2.1%) | 38 (2.1%) | 5 (1.7%) |  |

<sup>†</sup> Any TB symptom: cough, or weight loss, or fever, or weight loss.
<sup>‡</sup> Cough of 14 days or longer
\* Only recorded if patient had cough

HIV positive participants were more likely than HIV-negative or status-unknown participants to be female (72.9% vs 62.7% and 50.7%, p<0.001) and older (Median age 36 years vs 27 years and 27.5 years for HIV-positive, HIV-negative and HIV-unknown respectively, p<0.001) (Table 1). PLHIV were also more likely to be taking TB treatment (14.9% vs 3.2% and 1.8%), on IPT (21.5% vs 1.1% and 1.8%) and to report previous TB (22.6% vs 3.6% and 3.0% for HIV-positive, HIV-negative and HIV-unknown respectively) (all p<0.001). A higher proportion of PLHIV had chronic cough (15.1%) compared to HIV-negative (8.2%) or unknown-status participants (9.5%, p=0.001).

### TB diagnosis cascades

Overall 1,371 (57.2%) participants reported having been directly asked about coughing, with 128/2,397 (5.3% of total) asked to submit a sputum sample, 53/2,397 (2.2%) providing same-day sputum and 3/2,397 (0.13%) receiving same-day sputum results.

Diagnosis care cascades were constructed for clinically-indicated groups: HIV-negative participants with chronic cough, and PLHIV with any TB symptom (Figure 1). Proportions progressing through the sputum request and submission steps of the diagnostic pathway were higher for these key groups than other participants. In the 149 HIV-negative participants with chronic cough, 63.1% were asked about cough, 10.1% were asked for sputum, 6.7% gave sputum and none received same-day results. Among the 181 PLHIV with any TB symptom 60.2% were asked about cough, 13.3% were asked for sputum, 9.4% gave sputum and 0.6% received same-day results. Overall sputum submission for TB testing was achieved in 8.2% (27/330) of clinically-indicated participants.

**Figure 1:**
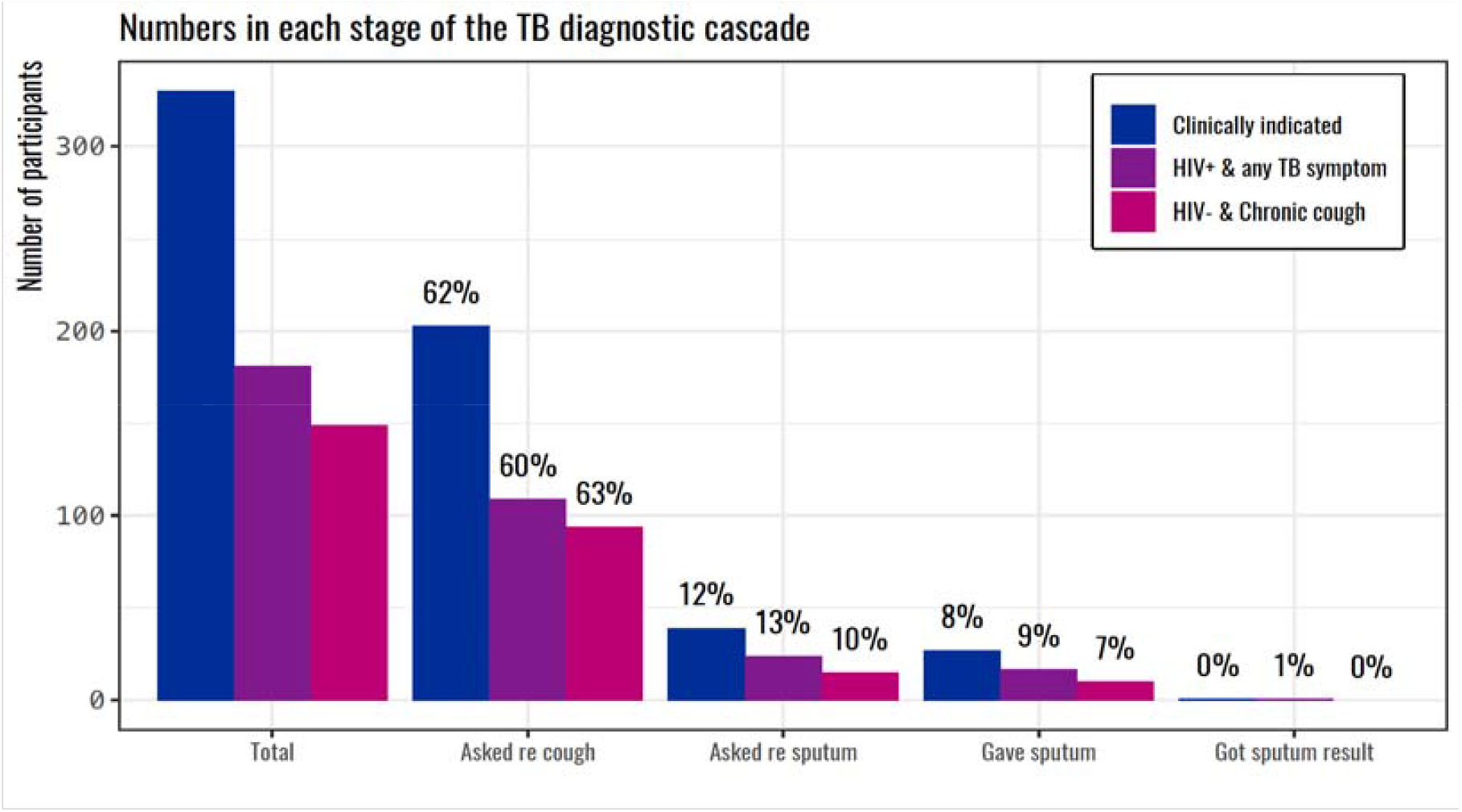
Diagnosis care cascades for groups clinically indicated for TB sputum tests.

Participants were lost at every step of the diagnosis cascade: 38.5% (127/330) of clinically indicated participants were not asked about cough, 49.7% (164/330) were then not asked for sputum despite having symptoms elicited and 3.6% (12/330) did not give sputum despite health worker request. For all clinically-indicated groups, the biggest gap in the diagnosis cascade was between symptom enquiry and requesting sputum. For HIV-negative participants with chronic cough, clinicians requested sputum for 13.8% (13/94) of those they had asked about cough and in PLHIV with any TB symptom this was 20.2% (22/109) (two people in both clinically-indicated groups were requested to give sputum but had not been asked about cough).

### Factors associated with being asked to submit sputum

On univariable analysis for all participants (Table 2), factors significantly associated with being asked to submit sputum included: older age (OR: 1.02, 95%CI: 1.01-1.03 per year increase in age), previous TB treatment (OR: 2.13, 95%CI: 1.08-4.20); being HIV-positive (OR: 1.69, 95%CI: 1.02-2.80); and presence of all TB symptoms (all p<0.001, except night sweats p=0.003).

**Table 2:** Univariable and multivariable associations with being asked to submit sputum: all participants.

| Variable | Unadjusted OR |  | Adjusted OR:<br>Any TB symptom |  | Adjusted OR:<br>Individual symptoms |  |
| --- | --- | --- | --- | --- | --- | --- |
|  | OR (95%<br>CI) | P value | aOR (95%<br>CI) | P value | aOR (95%<br>CI) | P value |
| Sex | 1.02<br>(0.69-1.49) | 0.936 | 1.01<br>(0.68-1.49) | 0.975 | 1.08<br>(0.73-1.62) | 0.695 |
| Age | 1.02<br>(1.01-1.03) | <0.001 | 1.02<br>(1.01-1.04) | <0.001 | 1.02<br>(1.01-1.03) | 0.002 |
| Previous TB | 2.13<br>(1.08-4.20) | 0.026 | 1.64<br>(0.79-3.37) | 0.183 | 1.59<br>(0.75-3.37) | 0.224 |
| HIV+* | 1.69<br>(1.02-2.80) | 0.040 | 1.42<br>(0.84-2.42) | 0.191 | 1.45<br>(0.85-2.49) | 0.174 |
| Any TB<br>symptom† | 3.27<br>(2.07-5.18) | <0.001 | 3.20<br>(2.02-5.06) | <0.001 | - | - |
| Cough <2<br>weeks | 2.48<br>(1.70-3.61) | <0.001 | - | - | 3.43<br>(2.23-5.28) | <0.001 |
| Chronic cough‡ | 3.32<br>(2.07-5.33) | <0.001 | - | - | 3.71<br>(2.10-6.56) | <0.001 |
| Weight loss | 2.52<br>(1.63-3.89) | <0.001 | - | - | 1.54<br>(0.96-2.47) | 0.076 |
| Fever | 2.10<br>(1.44-3.06) | <0.001 | - | - | 1.43<br>(0.94-2.18) | 0.096 |
| Night sweats | 1.86<br>(1.23-2.80) | 0.003 | - | - | 1.05<br>(0.66-1.68) | 0.827 |

\* Reference group: HIV-negative. Status unknown not presented † Any TB symptom: cough, or weight loss, or fever, or weight loss. ‡ Cough of 14 days or longer

On multivariable analysis increasing age (adjusted OR: 1.02, 95%CI: 1.01-1.04 per year) and any TB symptom (adjusted OR: 3.20, 95%CI: 2.02-5.06) or presence of cough (cough<2 weeks adjusted OR 3.43, 95%CI: 2.23-5.28, chronic (≥2 weeks) cough adjusted OR: 3.71, 95%CI: 2.10-6.56) remained significantly associated with being asked to submit sputum for all participants (Table 2).

On stratification by HIV status all these factors remained significantly associated for HIV-negative participants, but only the presence of any TB symptom (OR: 8.24, 95%CI: 1.08-37.68, adjusted OR: 8.18, 95%CI: 1.85-36.21) and chronic cough (OR: 10.84, 95%CI: 3.66-32.09, adjusted OR: 13.06, 95%CI: 3.69-46.28) were significantly associated with request for sputum amongst PLHIV (Table 3).

**Table 3:** Univariable and multivariable associations with being asked to submit sputum by HIV status.

|  | HIV-positive |  |  |  | HIV-negative |  |  |  |
| --- | --- | --- | --- | --- | --- | --- | --- | --- |
|  | Univariable<br>OR (95%<br>CI) | P value | Multivariable<br>aOR (95%<br>CI) | P value | Univariable<br>OR (95%<br>CI) | P value | Multivariable<br>aOR (95%<br>CI) | P value |
| Sex | 1.15<br>(0.40-3.29) | 0.801 | 1.08<br>(0.35-3.32) | 0.891 | 1.04<br>(0.67-1.62) | 0.867 | 1.18<br>(0.74-1.87) | 0.485 |
| Age | 1.02<br>(0.98-1.06) | 0.394 | 1.02<br>(0.98-1.06) | 0.383 | 1.02<br>(1.01-1.04) | 0.001 | 1.02<br>(1.01-1.04) | 0.008 |
| Previous<br>TB | 0.51<br>(0.11-2.28) | 0.367 | 0.52<br>(0.11-2.48) | 0.413 | 3.66<br>(1.67-8.06) | 0.001 | 3.37<br>(1.45-7.81) | 0.005 |
| Any TB<br>symptom† | 8.24<br>(1.80-37.68) | 0.001 | 8.18<br>(1.85-36.21) | 0.006 | 2.56<br>(1.56-4.20) | <0.001 | -* | -* |
| Cough<br><2 weeks | 1.28<br>(0.44-3.72) | 0.645 | -* | -* | 2.59<br>(1.68-4.01) | <0.001 | 3.16<br>(1.94-5.13) | <0.001 |
| Chronic<br>cough‡ | 10.84<br>(3.66-32.09) | <0.001 | -* | -* | 2.37<br>(1.30-4.33) | 0.004 | 2.56<br>(1.25-5.25) | 0.010 |

† Any TB symptom: cough, or weight loss, or fever, or weight loss.
‡ Cough of 14 days or longer
\* Multivariable analysis for HIV+ presented for Any TB symptom model, for HIV- presented model includes individual symptoms. Other symptoms (weight loss, fever and night sweats included in model but not presented: no significant relationship on multivariate analysis)

### Sputum test throughput and capacity

If all patients clinically indicated for a TB test did submit sputum (330/44%=750 over 78 working days) that would result in ∼20 sputum samples on each working day (10 patients a day, each with two samples). The clinic laboratory has one GeneXpert machine to process TB samples, with a maximum throughput of the of 8-12 samples a day (4 samples per cartridge with 2 hour run time plus preparation).

## Discussion

This study found that same day sputum submission for TB testing was achieved for only 8.2% of those where sputum testing was indicated according to Malawi national guidelines, with patients lost at every stage of the TB diagnosis care cascade. Failure to request sputum by clinicians despite elicited symptoms led to the biggest single gap in the diagnosis care cascade, followed by not asking about symptoms. This suggests that: interventions focusing on health worker behaviour may have the greatest potential for retaining presumptive TB patients within the diagnosis cascade; there appears to be inconsistent application of infection control practices; and that we must formalise and strengthen reporting on the early steps in the TB care cascade. Additional important epidemiological groups such as men should be given equal priority to PLHIV within national TB guidelines. However, if guideline adherence is improved, novel high-throughput triage testing approaches will also be needed to reach the required capacity.

Adherence to sputum-request guidelines in 11.8% (39/330) of patients is similar to that observed in India (12-17%) (15) and Uganda 13.2% (24) and sits at the lower end of the range (4-84%) identified in a recent systematic review (17). When taken together with a TB treatment initiation rate of 85-94% (25) and TB treatment success rate of 82% in Malawi (21), our data suggests that the overall TB cascade in Malawi is more similar to that for India than that for South Africa. In India gap 1 (“did not access a TB diagnostic test”) accounted for 50% of all patient losses, whereas in South Africa, low treatment success led to the largest gap in the cascade (8).

To reduce these substantial gaps in accessing TB tests a multi-faceted approach is required to identify logistical barriers and change health worker behaviours. Facility-based screening relies on health worker behaviour (asking about symptoms and requesting sputum) which leads to the biggest gaps and therefore offers the greatest potential for improvement. Suspicion of malaria or bacterial investigations may contribute to not requesting sputum (24) but further investigation is needed to confirm what drives health worker behaviour.

This study demonstrates a low level of adherence to National TB Programme guidelines. This is the case even with groups identified as high risk within both the Malawi and WHO guidelines, such as those who have previously had TB and PLHIV. Health workers operate in challenging conditions with average patient consultation times <3 minutes (26), a high turnover of staff and regular supply stock outs (27). As such, measures undertaken to improve adherence to guidelines and increase the proportion of clinically-indicated patients who access TB tests need to be pragmatic. Strategies such as FAST - Finding TB cases Actively, Separating safely and Treating effectively – (28) are effective in increasing testing and infection control not only for TB but also other respiratory infections. In Malawi, some elements of FAST, such as cough monitors, have been inconsistently implemented, due to limited availability of resources. However, our analysis shows the large gap in cough and symptom enquiry that could be met by universal cough monitors. Implementing FAST consistently is critical for all LMICs, especially in the midst of the COVID-19 global pandemic.

In addition, enhanced monitoring and central collation of data are essential to tracking individual clinic performance. Malawi, as is typical for LMICs, collects and reports comprehensive data on TB case notification and treatment success at clinic level, but only reports the number of TB tests per facility per quarter, without further diagnostic steps. A WHO requirement to report numbers of screened presumptive TB cases, disaggregated by age, gender and HIV-status globally, would allow greater focus on the earlier steps of the TB care cascade.

Despite TB prevalence in men being over twice as high as among women in LMICs (29) and in Malawi a ratio of male to female cases of 1.5 (21), in our study sex was not associated with being requested to submit sputum. In Malawi, the ratio of prevalent-to-notified cases of TB – an indication of how long patients take to be diagnosed - is 1.5 times higher among men than women (29). Men should, therefore, be considered as much of a priority group within TB guidelines as PLHIV in countries with a high male-to-female case ratio. Notably, men are less likely than women to seek health care early on in their illness (30), making it critical to manage them efficiently when they do present to a facility.

Finally, if all patients attending the outpatient clinic were screened for TB as per the guidelines, the current Xpert facilities would only be able to process up to two thirds of the required samples. If guideline adherence and increased identification of presumptive TB patients is subsequently improved a novel high-throughput approach to triage testing using new diagnostics (e.g. computer aided diagnostics for X-rays) will also be required for LMICs to increase capacity (31, 32).

Study limitations include the single site nature of this study, limiting generalisability. Due to limited research staff capacity we interviewed only 44% of clinic attendees, potentially resulting in selection bias, although this is mitigated by high participation in those approached. Symptoms, HIV status and testing practices were self-reported, potentially resulting in social desirability bias in measurement of these variables.

## Conclusion

Same-day sputum submission for TB testing was achieved in only 8.2% of those clinically indicated. Requesting sputum after eliciting symptoms is the key point of the cascade to intervene. Interventions are needed to optimise TB screening guidelines, formalise reporting, increase guideline adherence and move TB diagnosis away from an over-reliance on sputum testing, in order to reduce the most significant gaps early in the TB care cascade and to reach the required testing capacity to meet the WHO End TB goals.

## Data Availability

Data and code to reproduce this analysis is available from https://github.com/petermacp/tbcascade.

## Notes

### Competing Interest Statement

The authors have declared no competing interest.

### Funding Statement

Study was funded by the Wellcome Trust.
PM is funded by Wellcome (206575)
ELW received salary funding from the UK Medical Research Council (grant number MR/K012126/1), this award is jointly funded by the UK Medical Research Council (MRC) and the UK Department for International Development (DFID) under the MRC/DFID Concordat agreement and is also part of the EDCTP2 programme supported by the European Union.

### Author Declarations

The study was approved by the ethics committees of the Liverpool School of Tropical Medicine (17-050RS) and the College of Medicine, University of Malawi (P.11/17/2311).

